# Associations between e-cigarette use and e-cigarette flavors with cigarette smoking quit attempts and quit success: Evidence from a US large, nationally representative 2018-2019 survey

**DOI:** 10.1101/2022.03.31.22273144

**Authors:** Yoonseo Mok, Jihyoun Jeon, David T Levy, Rafael Meza

**Affiliations:** Department of Epidemiology, University of Michigan, Ann Arbor, MI; Department of Oncology, Georgetown University, Washington, DC

## Abstract

**Objectives:** While many studies have examined the association between e-cigarette use and smoking cessation, fewer have considered the impact of e-cigarette flavors on cessation outcomes. This study extends previous studies by examining the effects of e-cigarette use and e-cigarette flavors on smoking quit attempts and quit success.

**Methods:** We used data from the 2018-2019 Tobacco Use Supplement-Current Population Survey (TUS-CPS) survey. Multivariate logistic regression analyses were used to investigate the associations between e-cigarette and flavor use with quit attempts among individuals who smoked 12 months ago and quit success. Two current e-cigarette use definitions were considered; currently use every day or some days vs. 20+ days in the past 30-days.

**Results:** Compared to those not using e-cigarettes, current everyday or someday e-cigarette use with all non-tobacco flavors had an adjusted odds ratio (AOR) of 2.9 (95% CI: 2.4-3.5) for quit attempts and 1.7 (95% CI: 1.3-2.2) for quit success. 20+ days e-cigarette use with flavors had stronger associations with quit attempts (AOR=4.2, 95% CI: 3.1-5.5) and quit success (AOR=4.0, 95% CI: 2.9-5.4). E-cigarette users with non-tobacco flavors were more likely to succeed in quitting compared to those exclusively using non-flavored or tobacco-flavored e-cigarettes. Menthol/mint flavor users had slightly higher odds of quit attempts and success than users of other non-tobacco flavors.

**Conclusions:** E-cigarette use is positively associated with both making a smoking quit attempt and quit success. Those using flavored e-cigarettes, particularly menthol/mint, are more likely to quit successfully.

**Implications:** E-cigarette use is positively associated with both making a quit attempt and quit success, and those using flavored e-cigarettes are more likely to successfully quit smoking, with no statistically significant differences between use of menthol or mint flavored e-cigarettes versus use of other non-tobacco flavored products. This suggests that the potential for e-cigarettes to help people who currently smoke quit could be maintained with the availability of menthol or mint flavored e-cigarettes, even if other non-tobacco flavored products, which are associated with e-cigarette use among youth, were removed from the market.

## Introduction

Cigarette smoking remains the principal preventable cause of death in the US, with more than 480,000 smoking-attributable deaths per year.^1^ Thus, smoking prevention interventions and strategies to facilitate cessation for people who smoke remains a public health priority.

In 2006, e-cigarettes were first introduced to the US market, and their use has grown, resulting in an ongoing debate on their safety and regulation.^3^ Most e-cigarette users, particularly at older ages, currently smoke and use e-cigarettes either to quit smoking or just recreationally with no intention to quit, or formerly smoked.^4^ Some argue that dual use of cigarettes and e-cigarettes may reduce the concerns about health-related harms by those who smoke cigarettes, leading to extended smoking, and that e-cigarette use might act as a gateway to smoking, especially among youth.^5^ On the other hand, the potential of e-cigarettes to serve as a harm-reducing replacement for cigarettes or as an effective smoking cessation aid has been shown in some randomized control trials.^6^ In addition, some observational studies^7-14^ also indicate that e-cigarette use is associated with cessation behaviors and with higher rates of quit attempts and quit success, while others find conflicting results.^15-17^

In this study, we extended and adapted the approach by Levy et al.^8^ to examine the role of e-cigarettes in smoking quit attempts and quit success (remaining quitting smoking for at least 3 months). Like that study, we use TUS-CPS data, which includes specific information on quit attempts in the last year for people who smoke at the time of the survey, and the time since quitting for people who previously smoked. Unlike the Levy et al. study which used the TUS-CPS 2014-2015 data, we apply the more recent TUS-CPS 2018-2019 data, which provides information on different flavors among current e-cigarette users. Thereby, we consider more recent e-cigarette use during the period when flavored pod-based devices emerged. While many studies have examined the association between frequency of e-cigarette use and smoking cessation, few studies^12,18,19^ have considered the role of e-cigarette flavors on cessation.

## Methods

### Study population

We conducted a cross-sectional analysis of the relationship between e-cigarette use and cigarette smoking quit attempts and quit success using data from the Tobacco Use Supplement to the Current Population Survey (TUS-CPS) 2018-2019. TUS-CPS is a nationally representative survey administered as part of the U.S Census Bureau’s Current Population Survey. The TUS-CPS 2018-2019 data includes three samples collected in July 2018, January 2019, and May 2019. We limit the analysis to self-respondents, who were asked more detailed tobacco use questions and to those who were ages 18 and above. In addition, in order to analyze quitting behavior within the last year, the study population is restricted to individuals who reported smoking 12 months ago.

### Socio-demographic variables

Participants reported socio-demographic information including gender, age (18-21, 22-25, 26-29, 30-34, 35-44, 45-64, 65 years old and above), race (White, Black, Asian, Other Races), Hispanic origin (Hispanic/Non-Hispanic), education (less than 12^th^ grade, high school degree, some college but no degree, college graduate and above), family income levels (less than $19,999, $20,000-$39,999, $40,000-$74,999, $75,000 or more), marital status (never married, married-spouse present, married-spouse absent, or widowed/divorced/separated), employment status (employed/not in labor force or unemployed), residency status (metropolitan/non-metropolitan), and indoor work (yes/no).

### Cigarette smoking variables and cessation outcomes

All respondents in TUS-CPS were asked if they had smoked at least 100 cigarettes during their life and then if they now smoked cigarettes every day, some days, or not at all. Among the individuals with valid responses to these two questions (excluding “don’t know”, “refused”, “no response”), those who currently smoke were defined as individuals who had smoked at least 100 cigarettes in their lifetime and were smoking every or some days at the time of the survey.

People who currently smoke were then asked if they were “smoking cigarettes every day, some days, or not at all around this time 12 months ago.” The sample of people who smoked 12 months ago included those who currently smoke and reported smoking “every” or “some” days 12 months ago.

People who used to smoke and quit within 12 months were defined as individuals who had smoked at least 100 cigarettes in their lifetime and reported smoking 12 months ago but were not currently smoking at the time of the survey. The sample of people who smoked 12 months ago also included these individuals.

There was a total sample of 17,205 individuals who smoked 12 months ago, of which 15,049 currently smoked at the time of the survey and 2,156 quit within the past 12 months. Among those, people who currently smoke with unknown quit attempts, and people who used to smoke and quit within the last 12 months but reported no cigarettes smoked 12 months ago or unknown smoking frequency 12 months ago were omitted, leaving a study sample of 16,591 (Figure S1).

People who used to smoke 12 months ago were categorized by the number of cigarettes smoked per day (cpd): very light (fewer than 5 cpd), light (5-14 cpd), medium (15-24 cpd), and heavy (25 or more cpd). People who reported smoking daily 12 months ago, were asked “the average number of cigarettes smoked per day 12 months ago.” For those who smoked non-daily 12 months ago, their reported number of cigarettes per day on the days they smoked were multiplied by the number of days smoked per month and divided by 30 to measure average cpd use. An indicator variable was also included for those who classified themselves as “some days” cigarette users.

Individuals who used to smoke and quit within 12 months were grouped into categories based on their time since quit; quitting 3-12 months, 1 to less than 3 months, and less than 1 month ago.

#### Quit attempts and quit success

The respondents who smoked 12 days or less in the past 30-days around this time 12 months ago were asked whether they had “tried to quit smoking completely during the past 12 months.” Respondents who smoked more than 12 days were asked whether they “stopped smoking for one day or longer because of trying to quit smoking during the past 12 months.” People who smoked 12 months ago were considered to have made a quit attempt if they answered “yes” to either of these questions.

### E-cigarette use, e-cigarette flavors, and smokeless tobacco use

Three questions were asked about current and past e-cigarette use. After a description of e-cigarettes, participants were asked, “Have you ever used e-cigarettes even one time?” Participants were classified as ever users of e-cigarettes if they answered “yes” to this question. Ever users of e-cigarettes were then asked, “Do you now use an e-cigarette every day, some days, or not at all?”. Current e-cigarette users include those who answered “every day” or “some days.” Non-users include those who do not currently use “every day” or “some days.” Those who responded “some days” were then asked, “On how many of the past 30-days did you use e-cigarettes?” Among current users, we differentiated those who currently used e-cigarettes at least 20 days in the past 30-days, based on results in Levy et al. An equivalent measure was also developed for smokeless tobacco (SLT) use.

We also classified e-cigarette use by flavors. E-cigarette flavors were assessed in two questions. Current e-cigarette users were asked whether they usually used flavored e-cigarettes and to indicate which of the 4 flavor categories they used (select all that apply: “Tobacco,” “Menthol or mint,” “Fruit, candy, sweets, chocolate, clove, spice, herb, or alcohol,” “Other”). Respondents indicating not using flavored e-cigarettes were further asked whether they usually used tobacco-flavored e-cigarettes. We created two e-cigarette use variables distinguishing flavor use. The first variable categorized individuals as “non-e-cigarette users”, “currently use non-flavored or exclusive tobacco-flavored e-cigarettes” or “currently use flavored e-cigarettes, except tobacco-flavored”. For the second variable, we further stratified the “currently use flavored e-cigarettes, except tobacco-flavored” category into “currently use menthol or mint-flavored e-cigarettes regardless of using other flavors” and “currently use flavored e-cigarettes but not tobacco-flavored nor menthol or mint flavored.”

### Statistical Methods

For each outcome (quit attempts and quit success), chi-square analyses were conducted to test the differences within category for each of the socio-demographic variables, smoking frequency, measures of e-cigarette use, e-cigarette flavors, and SLT use.

Separate multivariate logistic regression models were fit to investigate the association of e-cigarette use and either quit attempts or quit success. In the quit attempts model, the sample included those who smoked 12 months ago, and the outcome was whether those individuals made a quit attempt in the last year. In the quit success model, the sample was limited to those having made a quit attempt, distinguishing those who failed and those who quit for longer than 3 months. Only individuals who used to smoke and who remained quit for at least 3 months were considered as quitting successfully to capture those who are more likely to remain abstinent among those who have made a quit attempt (Figure 1). Consistent with Levy et al, individuals who used to smoke with less than three months since quitting were removed from this analysis.^8^

**Figure 1.**
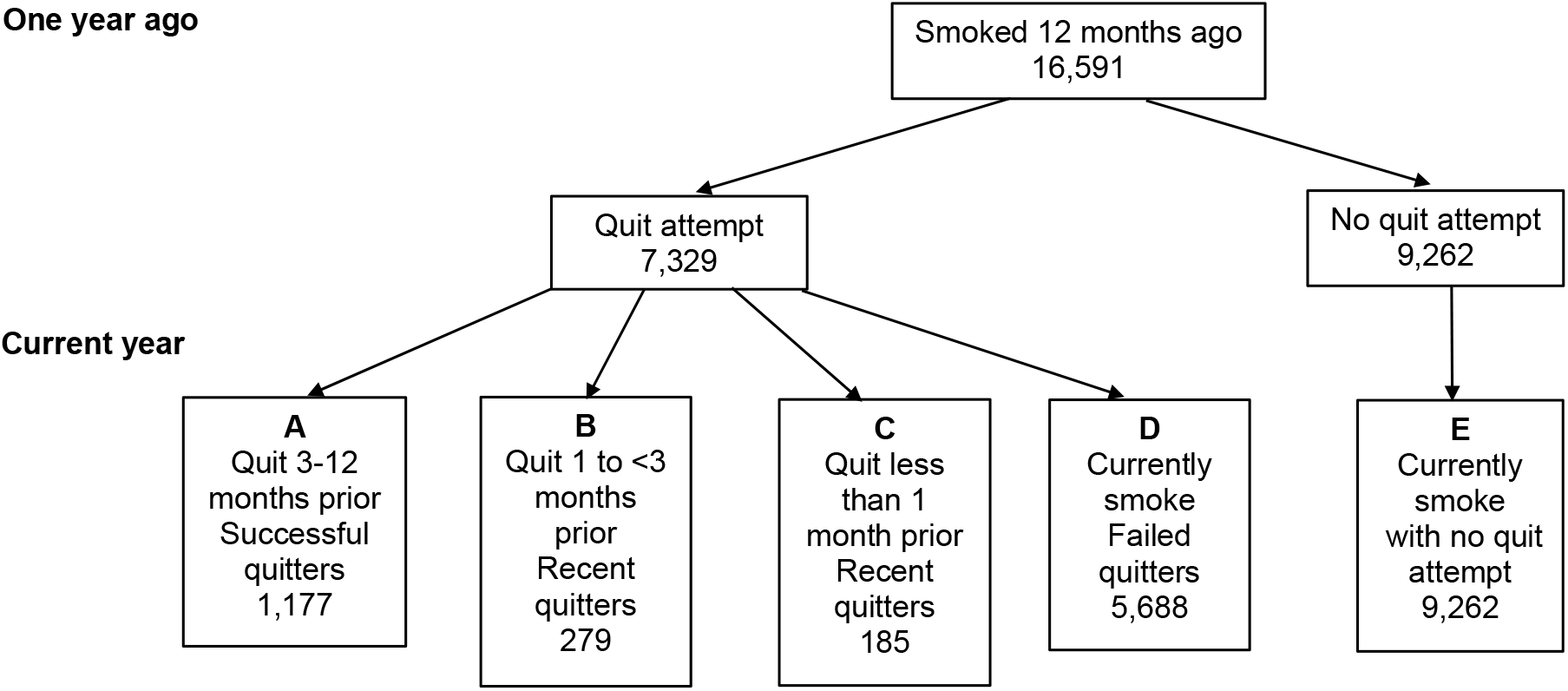
Sample design.

We computed adjusted odds ratios (AORs) for quit attempts or quit success by including all the other covariates in the analyses. Multivariate models were fit using the “survey” package logistic regression in the R statistical program (version 4.0). All estimates accounted for self-response sample weights.

## Results

### Descriptive Statistics: Quit Attempts and Quit Success Rates

The rate (percentage %) of people who smoked 12 months ago and made a quit attempt, and the rate of quit success among those who made a quit attempt within the last 12 months are presented in Table 1. The rate of making quit attempts significantly differed by all individual characteristics except employment status and SLT use. Individuals with lower cigarettes per day (cpd) had a higher quit attempt rate: 1-4 (55.6%), 5-14 (46.1%), 15-24 (38.0%), 25+ (33.3%). Quit attempts significantly differed by the e-cigarette use and flavor: “currently use non-flavored or exclusive tobacco-flavored e-cigarettes” (59.0%), “currently use flavored e-cigarettes, except tobacco-flavored” (69.9%), “non-e-cigarette users” (42.1%).

**Table 1.** Quit attempts among people who smoked 12 months ago and quit success among those making a quit attempt

| Variable | Categories | Quit attempt model |  |  | Quit success model |  |  |
| --- | --- | --- | --- | --- | --- | --- | --- |
|  |  | Sample size | Quit attempts | Chi-square | Sample Size | Quit success | Chi-square |
|  |  |  | % | (p value) |  | % | (p value) |
| Overall |  | 16591 | 44.2% |  | 6865 | 17.1% |  |
| Gender | Male | 8368 | 42.6% | 17.83 | 3324 | 17.7% | 1.13 |
|  | Female | 8223 | 45.8% | (<0.001) | 3541 | 16.7% | (0.287) |
| Age | 18-21 | 273 | 57.5% |  | 141 | 25.5% |  |
|  | 22-25 | 692 | 59.3% |  | 373 | 26.0% |  |
|  | 26-29 | 1058 | 55.3% |  | 544 | 26.7% |  |
|  | 30-34 | 1602 | 49.4% |  | 728 | 20.2% |  |
|  | 35-44 | 3100 | 43.3% |  | 1263 | 17.7% |  |
|  | 45-64 | 7093 | 41.2% | 196.01 | 2765 | 12.8% | 104.65 |
|  | 65+ | 2773 | 40.5% | (<0.001) | 1051 | 16.8% | (<0.001) |
| Race | White | 13768 | 43.3% |  | 5563 | 18.2% |  |
|  | Black | 1772 | 48.6% |  | 827 | 11.1% |  |
|  | Asian | 344 | 47.7% | 26.75 | 142 | 17.6% | 26.65 |
|  | Other Races | 707 | 48.9% | (<0.001) | 333 | 14.7% | (<0.001) |
| Hispanic | Hispanic | 1169 | 47.4% | 5.14 | 527 | 19.0% | 1.21 |
|  | Non-Hispanic | 15422 | 43.9% | (0.023) | 6338 | 17.0% | (0.271) |
| Education | Less than 12<sup>th</sup> grade | 2335 | 40.9% |  | 913 | 11.4% |  |
|  | High school degree | 6479 | 41.9% |  | 2551 | 15.4% |  |
|  | Some college, no degree | 3588 | 47.1% | 50.26 | 1577 | 18.1% | 54.17 |
|  | College degree or higher | 4189 | 47.0% | (<0.001) | 1824 | 21.7% | (<0.001) |
| Family income | \$0-\$19999 | 4060 | 46.1% | | 1772 | 13.5% | |
| | \$20000-\$39999 | 4456 | 42.6% | | 1755 | 15.7% | |
| | \$40000-\$74999 | 4551 | 43.5% | 12.53 | 1866 | 17.9% | 47.27 |
| | \$75000 or more | 3524 | 44.8% | (0.006) | 1472 | 22.3% | (<0.001) |
| Marital status | Never Married | 4656 | 47.7% |  | 2070 | 18.9% |  |
|  | Married Present | 5900 | 42.1% |  | 2326 | 19.1% |  |
|  | Married-Spouse Absent | 275 | 41.8% | 34.82 | 109 | 18.4% | 31.86 |
|  | Widowed/Divorced/Separated | 5760 | 43.6% | (<0.001) | 2360 | 13.6% | (<0.001) |
| Employment | Employed | 9518 | 44.7% | 2.50 | 3970 | 18.8% | 18.79 |
|  | Not in labor force or unemployed | 7073 | 43.5% | (0.114) | 2895 | 14.8% | (<0.001) |
| Metropolitan status | Metropolitan | 11963 | 45.1% | 16.04 | 5069 | 18.0% | 8.67 |
|  | Non-metropolitan | 4628 | 41.7% | (<0.001) | 1796 | 14.9% | (0.003) |
| Indoor workers | No | 10559 | 43.0% | 15.44 | 4275 | 15.9% | 12.94 |
|  | Yes | 6032 | 46.2% | (<0.001) | 2590 | 19.3% | (<0.001) |
| Cigarettes per day 12 months ago | 1-4 | 3197 | 55.6% |  | 1682 | 20.5% |  |
|  | 5-14 | 5923 | 46.1% |  | 2552 | 14.5% |  |
|  | 15-24 | 5668 | 38.0% |  | 2007 | 17.8% |  |
|  | 25+ | 1259 | 33.3% | 325.84 | 385 | 21.0% | 39.84 |
|  | Unknown CPD | 544 | 45.0% | (<0.001) | 239 | 9.6% | (<0.001) |
| Smoking frequency | Smoked every day 12 months ago | 13226 | 40.4% | 382.50 | 4957 | 17.1% | 0.06 |
|  | Smoked some days 12 months ago | 3365 | 59.1% | (<0.001) | 1908 | 17.4% | (0.809) |
| Smokeless tobacco current use | Yes | 416 | 42.6% | 0.35 | 166 | 19.9% | 0.68 |
|  | No | 15963 | 44.1% | (0.557) | 6595 | 17.1% | (0.408) |
| E-cigarette | Current e-cig user | 1364 | 66.6% | 305.86 | 819 | 25.4% | 43.51 |
|  | Non e-cig user | 15012 | 42.0% | (<0.001) | 5943 | 16.1% | (<0.001) |
|  | Current e-cig user with non-flavored or exclusive tobacco-flavored | 410 | 59.0% |  | 228 | 20.6% |  |
|  | Current e-cig user with flavors | 954 | 69.9% | 320.65 | 591 | 27.2% | 49.24 |
|  | Non e-cig user | 15012 | 42.1% | (<0.001) | 5943 | 16.1% | (<0.001) |
|  | Current e-cig user with non-flavored or exclusive tobacco-flavored | 410 | 59.0% |  | 228 | 20.6% |  |
|  | Current e-cig user with menthol or mint flavor | 303 | 71.6% |  | 198 | 27.8% |  |
|  | Current e-cig user with other flavors | 651 | 69.1% | 321.17 | 393 | 27.0% | 49.30 |
|  | Non e-cig user | 15012 | 42.1% | (<0.001) | 5943 | 16.1% | (<0.001) |

The rate of quit success among those who made at least one quit attempt in the past 12 months showed significant differences by individual characteristics, except gender, Hispanic/Non-Hispanic, smoking frequency, and SLT use. There was a clear positive gradient in quit success rates by level of income or education, i.e., individuals with higher income or education levels had a higher rate of quit success. Similar to quit attempts, the rate of quit success significantly differed by e-cigarette use and flavor: 27.2% of “current users of flavored e-cigarettes, except tobacco-flavored” succeeded in quitting smoking, while 20.6% and 16.1% of “current users of non-flavored or exclusive tobacco-flavored” and “non-e-cigarette users” succeeded in quitting. No differences were observed when further stratifying the e-cigarette flavor categories (quit success rate of 27.8% for current e-cigarette users with menthol or mint flavors vs 27.0% for current e-cigarette users with other non-tobacco flavors).

### Logistic Regression Analysis: Quit Attempts

Table 2 presents the results for quit attempts. The first two columns show results when using an e-cigarette use definition of every day or some days. The last two columns show results when using a stricter e-cigarette use definition of at least 20 of the past 30-days. In both analyses, results indicate that females and those below age 35, Black or Other races, those with higher levels of education but lower levels of income were all more likely to make a quit attempt. Individuals who smoke some days, those who smoke fewer cigarettes per day and those who use e-cigarettes were also more likely to make a quit attempt.

**Table 2.**
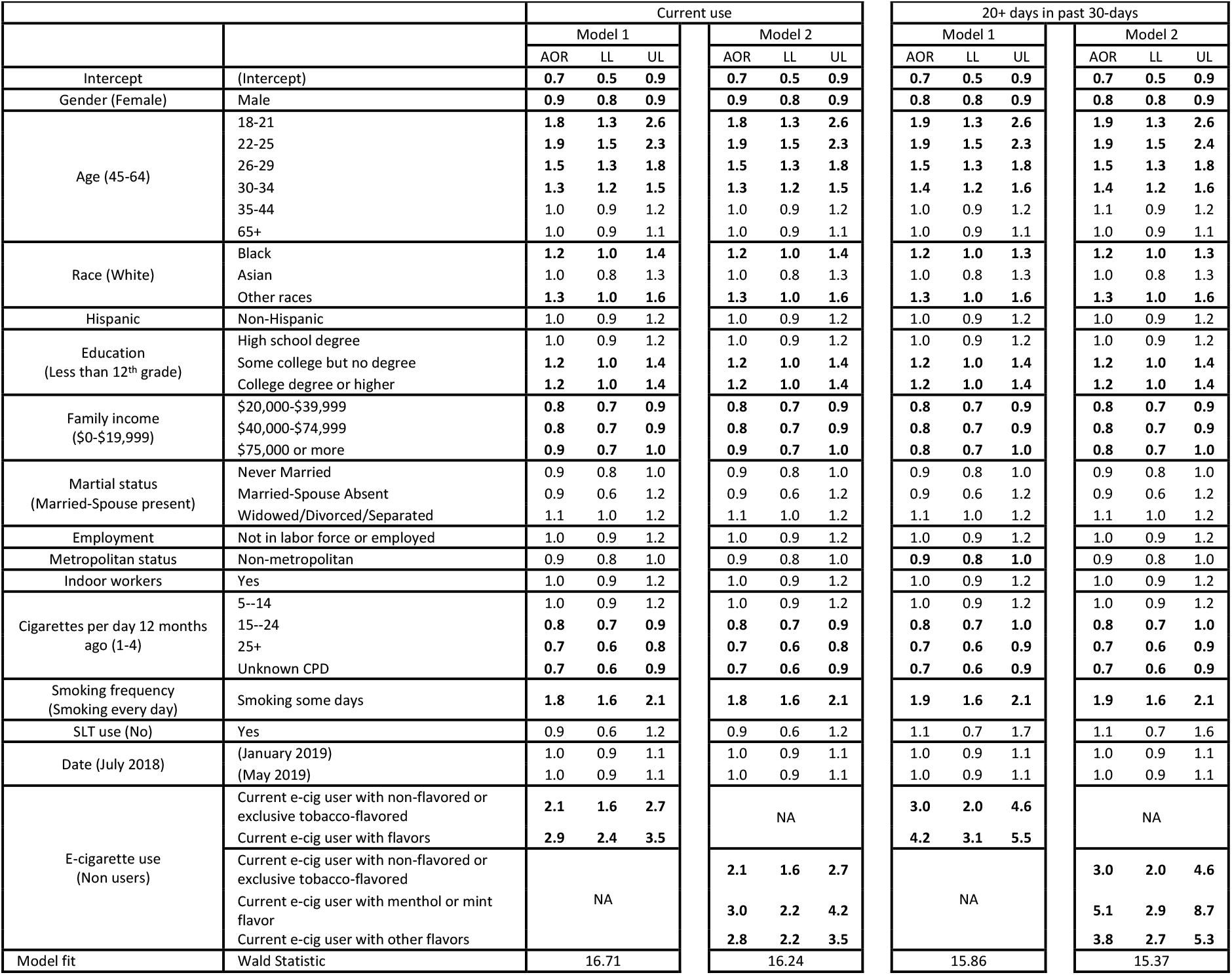
Logistic regression analysis of having made a quit attempt among individuals who smoked 12 months ago.

|  |  | Current use |  |  |  |  |  | 20+ days in past 30-days |  |  |  |  |  |
| --- | --- | --- | --- | --- | --- | --- | --- | --- | --- | --- | --- | --- | --- |
|  |  | Model 1 |  |  | Model 2 |  |  | Model 1 |  |  | Model 2 |  |  |
|  |  | AOR | LL | UL | AOR | LL | UL | AOR | LL | UL | AOR | LL | UL |
| Intercept | (Intercept) | 0.7 | 0.5 | 0.9 | 0.7 | 0.5 | 0.9 | 0.7 | 0.5 | 0.9 | 0.7 | 0.5 | 0.9 |
| Gender (Female) | Male | 0.9 | 0.8 | 0.9 | 0.9 | 0.8 | 0.9 | 0.8 | 0.8 | 0.9 | 0.8 | 0.8 | 0.9 |
| Age (45-64) | 18-21 | 1.8 | 1.3 | 2.6 | 1.8 | 1.3 | 2.6 | 1.9 | 1.3 | 2.6 | 1.9 | 1.3 | 2.6 |
|  | 22-25 | 1.9 | 1.5 | 2.3 | 1.9 | 1.5 | 2.3 | 1.9 | 1.5 | 2.3 | 1.9 | 1.5 | 2.4 |
|  | 26-29 | 1.5 | 1.3 | 1.8 | 1.5 | 1.3 | 1.8 | 1.5 | 1.3 | 1.8 | 1.5 | 1.3 | 1.8 |
|  | 30-34 | 1.3 | 1.2 | 1.5 | 1.3 | 1.2 | 1.5 | 1.4 | 1.2 | 1.6 | 1.4 | 1.2 | 1.6 |
|  | 35-44 | 1.0 | 0.9 | 1.2 | 1.0 | 0.9 | 1.2 | 1.0 | 0.9 | 1.2 | 1.1 | 0.9 | 1.2 |
|  | 65+ | 1.0 | 0.9 | 1.1 | 1.0 | 0.9 | 1.1 | 1.0 | 0.9 | 1.1 | 1.0 | 0.9 | 1.1 |
| Race (White) | Black | 1.2 | 1.0 | 1.4 | 1.2 | 1.0 | 1.4 | 1.2 | 1.0 | 1.3 | 1.2 | 1.0 | 1.3 |
|  | Asian | 1.0 | 0.8 | 1.3 | 1.0 | 0.8 | 1.3 | 1.0 | 0.8 | 1.3 | 1.0 | 0.8 | 1.3 |
|  | Other races | 1.3 | 1.0 | 1.6 | 1.3 | 1.0 | 1.6 | 1.3 | 1.0 | 1.6 | 1.3 | 1.0 | 1.6 |
| Hispanic | Non-Hispanic | 1.0 | 0.9 | 1.2 | 1.0 | 0.9 | 1.2 | 1.0 | 0.9 | 1.2 | 1.0 | 0.9 | 1.2 |
| Education<br>(Less than 12<sup>th</sup> grade) | High school degree | 1.0 | 0.9 | 1.2 | 1.0 | 0.9 | 1.2 | 1.0 | 0.9 | 1.2 | 1.0 | 0.9 | 1.2 |
|  | Some college but no degree | 1.2 | 1.0 | 1.4 | 1.2 | 1.0 | 1.4 | 1.2 | 1.0 | 1.4 | 1.2 | 1.0 | 1.4 |
|  | College degree or higher | 1.2 | 1.0 | 1.4 | 1.2 | 1.0 | 1.4 | 1.2 | 1.0 | 1.4 | 1.2 | 1.0 | 1.4 |
| Family income<br>(\$0-\$19,999) | \$20,000-\$39,999 | 0.8 | 0.7 | 0.9 | 0.8 | 0.7 | 0.9 | 0.8 | 0.7 | 0.9 | 0.8 | 0.7 | 0.9 |
| | \$40,000-\$74,999 | 0.8 | 0.7 | 0.9 | 0.8 | 0.7 | 0.9 | 0.8 | 0.7 | 0.9 | 0.8 | 0.7 | 0.9 |
| | \$75,000 or more | 0.9 | 0.7 | 1.0 | 0.9 | 0.7 | 1.0 | 0.8 | 0.7 | 1.0 | 0.8 | 0.7 | 1.0 |
| Marital status<br>(Married-Spouse present) | Never Married | 0.9 | 0.8 | 1.0 | 0.9 | 0.8 | 1.0 | 0.9 | 0.8 | 1.0 | 0.9 | 0.8 | 1.0 |
|  | Married-Spouse Absent | 0.9 | 0.6 | 1.2 | 0.9 | 0.6 | 1.2 | 0.9 | 0.6 | 1.2 | 0.9 | 0.6 | 1.2 |
|  | Widowed/Divorced/Separated | 1.1 | 1.0 | 1.2 | 1.1 | 1.0 | 1.2 | 1.1 | 1.0 | 1.2 | 1.1 | 1.0 | 1.2 |
| Employment | Not in labor force or employed | 1.0 | 0.9 | 1.2 | 1.0 | 0.9 | 1.2 | 1.0 | 0.9 | 1.2 | 1.0 | 0.9 | 1.2 |
| Metropolitan status | Non-metropolitan | 0.9 | 0.8 | 1.0 | 0.9 | 0.8 | 1.0 | 0.9 | 0.8 | 1.0 | 0.9 | 0.8 | 1.0 |
| Indoor workers | Yes | 1.0 | 0.9 | 1.2 | 1.0 | 0.9 | 1.2 | 1.0 | 0.9 | 1.2 | 1.0 | 0.9 | 1.2 |
| Cigarettes per day 12 months<br>ago (1-4) | 5--14 | 1.0 | 0.9 | 1.2 | 1.0 | 0.9 | 1.2 | 1.0 | 0.9 | 1.2 | 1.0 | 0.9 | 1.2 |
|  | 15--24 | 0.8 | 0.7 | 0.9 | 0.8 | 0.7 | 0.9 | 0.8 | 0.7 | 1.0 | 0.8 | 0.7 | 1.0 |
|  | 25+ | 0.7 | 0.6 | 0.8 | 0.7 | 0.6 | 0.8 | 0.7 | 0.6 | 0.9 | 0.7 | 0.6 | 0.9 |
|  | Unknown CPD | 0.7 | 0.6 | 0.9 | 0.7 | 0.6 | 0.9 | 0.7 | 0.6 | 0.9 | 0.7 | 0.6 | 0.9 |
| Smoking frequency<br>(Smoking every day) | Smoking some days | 1.8 | 1.6 | 2.1 | 1.8 | 1.6 | 2.1 | 1.9 | 1.6 | 2.1 | 1.9 | 1.6 | 2.1 |
| SLT use (No) | Yes | 0.9 | 0.6 | 1.2 | 0.9 | 0.6 | 1.2 | 1.1 | 0.7 | 1.7 | 1.1 | 0.7 | 1.6 |
| Date (July 2018) | (January 2019) | 1.0 | 0.9 | 1.1 | 1.0 | 0.9 | 1.1 | 1.0 | 0.9 | 1.1 | 1.0 | 0.9 | 1.1 |
|  | (May 2019) | 1.0 | 0.9 | 1.1 | 1.0 | 0.9 | 1.1 | 1.0 | 0.9 | 1.1 | 1.0 | 0.9 | 1.1 |
| E-cigarette use<br>(Non users) | Current e-cig user with non-flavored or<br>exclusive tobacco-flavored | 2.1 | 1.6 | 2.7 | NA |  |  | 3.0 | 2.0 | 4.6 | NA |  |  |
|  | Current e-cig user with flavors | 2.9 | 2.4 | 3.5 |  |  |  | 4.2 | 3.1 | 5.5 |  |  |  |
|  | Current e-cig user with non-flavored or<br>exclusive tobacco-flavored | NA |  |  | 2.1 | 1.6 | 2.7 | NA |  |  | 3.0 | 2.0 | 4.6 |
|  | Current e-cig user with menthol or mint<br>flavor |  |  |  | 3.0 | 2.2 | 4.2 |  |  |  | 5.1 | 2.9 | 8.7 |
|  | Current e-cig user with other flavors |  |  |  | 2.8 | 2.2 | 3.5 |  |  |  | 3.8 | 2.7 | 5.3 |
| Model fit | Wald Statistic | 16.71 |  |  | 16.24 |  |  | 15.86 |  |  | 15.37 |  |  |

Among e-cigarette users, two different flavor categorizations were considered. In the first, where any flavored e-cigarette use was distinguished from non-flavored or exclusive tobacco-flavored (model 1), both non-flavored or exclusive tobacco-flavored e-cigarette users (AOR 2.1, 95% CI: 1.6-2.7) and flavored e-cigarette users (AOR 2.9, 95% CI: 2.4-3.5) had significantly higher rates of quit attempts compared to non-e-cigarette users. When menthol or mint were further distinguished from other non-tobacco flavors (model 2), both flavored categories of e-cigarette users showed higher rates of quit attempts than non-flavored or exclusive tobacco-flavored e-cigarette users, although all e-cigarette use categories had statistically significant higher rates of quit attempts versus e-cigarette non-users. Interestingly, current e-cigarette users of menthol or mint flavors had slightly higher odds of making a quit attempt (AOR 3.0, 95% CI: 2.2-4.2) versus current e-cigarette users of other non-tobacco flavors (AOR 2.8, 95% CI: 2.2-3.5), although the difference was not statistically significant (model 2).

Similar results were seen when restricting e-cigarette use to those who used 20 of the past 30-days, but frequent users had higher rates of quit attempts (generally AOR of 3-5 compared to AOR of 2-3) than current e-cigarette everyday or someday users.

### Logistic Regression Analysis: Quit Success

Table 3 presents the results for quit success. In general, we find that those below age 45, with higher levels of education, higher levels of income and living in metropolitan areas were more likely to succeed in quitting smoking. Current everyday or someday e-cigarette users who use flavored e-cigarettes were more likely to quit smoking successfully compared to e-cigarette non-users (model 3, AOR 1.7, 95% CI: 1.3-2.2). However, e-cigarette users of non-flavored or exclusive tobacco-flavored products did not have higher quit success rates versus e-cigarette non-users (AOR 1.2, 95% CI: 0.8-1.8). When menthol or mint-flavored e-cigarette users were distinguished from users of e-cigarettes with other non-tobacco flavors, their likelihood of quitting smoking (AOR 1.9, 95% CI: 1.3.-2.9) was slightly higher than that of e-cigarette users with other non-tobacco flavors (AOR 1.6, 95% CI: 1.2-2.2), although the difference was not statistically significant (model 4). When considering only frequent e-cigarette use (20 +days), e-cigarette users had higher rates of quit success versus non-users regardless of flavor use (models 3 & 4).

**Table 3.**
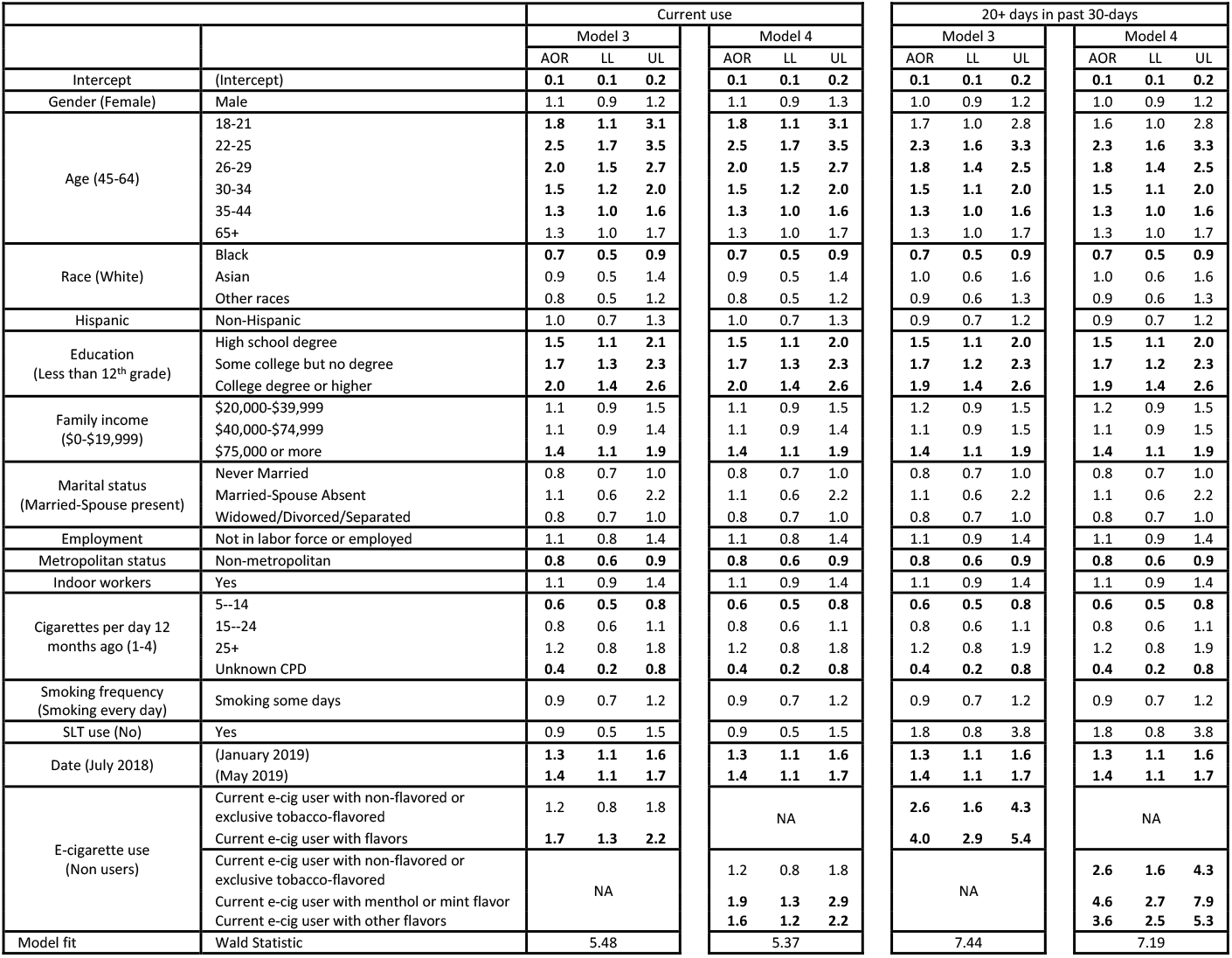
Logistic regression analysis of having made a quit success among individuals who smoked 12 months ago and made at least one quit attempt

## Discussion

The results clearly indicate that those who use e-cigarettes more intensely (at least 20 of the past 30-days) and those who use flavored e-cigarettes have both a higher odds of making a quit attempt and of succeeding in quitting cigarette smoking. Current e-cigarette users of menthol or mint flavors had higher odds of making quit attempts and quit success versus current e-cigarette users of other non-tobacco flavors, although the differences were not statistically significant.

The consistency of our findings with results from randomized control trials of e-cigarettes as smoking cessation aids^6^ and with those from other observational studies strengthens the evidence that e-cigarettes can help people who smoke quit.^7-14^ In particular, our results are consistent with previous studies using the earlier TUS-CPS 2014-2015 data that reported associations of e-cigarette use, particularly frequent use, with smoking cessation.^7,8^ Our findings are also consistent with another study of TUS-CPS 2018-2019 data that found that e-cigarette use is associated with smoking cessation contemplation and preparation, with stronger associations with more frequent e-cigarette use.^12^ Studies of the nationally-representative longitudinal Population Assessment of Tobacco and Health (PATH) data have also reported associations of e-cigarette use with smoking abstinence^20^ and with smoking cessation.^13,14^ In contrast, some other PATH studies have found no relationship of e-cigarette use with increased smoking cessation^15^ or reduced smoking relapse.^16,17^ Further research, particularly studies covering more recent periods with newer generation e-cigarette products, are needed to reconcile these differences.

Consistent with our results that flavored e-cigarette use is associated with increased odds of making a cigarette smoking quit attempt and quit success, using waves 1 to 4 of the PATH data, Friedman et al.^18^ found that flavored e-cigarettes use is associated with smoking cessation in adults. In addition, a cross-sectional study using data of adults from Canada and the U.S.^19^ also found increase odds of making a quit attempt when using flavored e-cigarettes. This study also found that non-tobacco flavors (menthol/mint, fruit, candy, or other) were most likely to be used among those who used to smoke, which is consistent with our findings of the association of flavored e-cigarette use with quit success. In contrast with these findings and ours, a recent analysis of the association of e-cigarette use with cessation behaviors in TUS-CPS 2018-2019 found no association between e-cigarettes with non-tobacco flavors and with contemplation or preparation to quit relative to use of tobacco-flavored e-cigarettes.^12^ Further studies are thus needed to better understand the role that e-cigarette flavors can have in different aspects of smoking cessation and relapse among recent quitters.

Our findings indicate that e-cigarette non-tobacco flavors can be helpful for smoking cessation. Interestingly, we found no evidence of a difference in the odds of quit attempts or success between users of menthol/mint versus other non-tobacco flavors. This suggests that the potential for e-cigarettes to help people who currently smoke quit could be maintained with the availability of menthol or mint flavored e-cigarettes, even if other non-tobacco flavored products, which are associated with e-cigarette use among youth,^21,22^ were removed from the market. However, the role of e-cigarettes in supporting smoking cessation could be somewhat diminished to the extent that some potential users might prefer sweetened to menthol or mint flavors.

Our study has some key limitations. Like other cross-sectional association studies, our results depend on the retrospective statements about behavior in the past year rather than observed behavioral changes as in longitudinal data. Related, e-cigarette flavor categorization was based on self-reported use at the time of the survey. However, e-cigarette users may have used flavored and non-flavored products at different periods during the past year and at different points in their quit attempt or quit success process. Further longitudinal observational and randomized smoking cessation studies of e-cigarettes and flavors use among people who currently smoke are thus needed to better assess their causal role in cessation outcomes. Another key limitation is that we did not distinguish e-cigarette use by device type or the nicotine strength of the liquids, which may be other key product features that influence quit attempt or quit success rates. Preliminary analyses (data not shown) evaluating the interactions of device type and flavors suggest that users of tank e-cigarettes who use flavored liquids have higher quit success rates than users of tank e-cigarettes with non-flavored or exclusive tobacco-flavored liquids.

Despite these limitations, our study indicates that e-cigarette use is positively associated with both making a quit attempt and quit success, and those using flavored e-cigarettes are more likely to successfully quit smoking, with no statistically significant differences between use of menthol or mint flavored e-cigarettes versus use of other non-tobacco flavored products. Future studies should investigate the relationship of frequency of e-cigarette use with smoking quit attempts or success using longitudinal data or conducting randomized controlled trials. In addition, further studies investigating the joint effects of e-cigarette device type, nicotine content and flavors on smoking cessation are needed. It will be also important to follow those who may have different patterns of e-cigarette use over time, and to consider the impact of newer versus older generations of e-cigarettes in helping smokers quit and in helping recent quitters avoid relapse.

## Data Availability

All data used is publicly available

