## Supplemental Figure 1 for "Associations between e-cigarette use and e-cigarette flavors with cigarette smoking quit attempts and quit success: Evidence from a US large, nationally representative 2018-2019 survey"

**Figure S1.** Flow chart of exclusions for the study sample

Self-respondents and 18 years or older (N=137,471)

Non-established smoking (N=91,442)

Now smoke “not at all” (N=29,189)

Has been less than or equal to 1 year since completely quitting smoking cigarettes (N=2,156)

Has been more than 1 year since completely quitting smoking cigarettes (N=27,033)

Unknown smoking frequency 12 months ago (N=38)

Smoked “every day” or “some days” 12 months ago (N=1,679)

Known smoking frequency 12 months ago (N=1,641)

Responded “yes” or “no” to have you ever tried to quit smoking? (N=14,950)

Unknown response to have you ever tried to quit smoking? (N=99)

Smoked “every day” or “some days” 12 months ago (N=15,049)

Not smoked 12 months ago (N=1,521)

Now smoke every day or some days (N=16,570)

Unknown current smoking status (N=270)

Known current smoking status (N=45,759)

Established smoking (N=46,029)

Total currently smoke (N=14,950)

Total formerly smoked (N=1,641)

Reported not smoking 12 months ago (N=477)
